# CD38 expression by antigen-specific CD4 T cells correlates with sputum bacterial load at time of tuberculosis diagnosis and is significantly restored 5-months after treatment initiation

**DOI:** 10.1101/2022.01.26.22269875

**Authors:** Hellen Hiza, Jerry Hella, Ainhoa Arbués, Mohamed Sasamalo, Veronica Misana, Jacques Fellay, Sébastien Gagneux, Klaus Reither, Damien Portevin

## Abstract

T cell activation markers (TAM) expressed by antigen-specific T cells constitute promising candidates to attest the presence of an active infection by *Mycobacterium tuberculosis* (*Mtb*). Reciprocally, their modulation may be used to assess antibiotic treatment efficacy and eventually attest disease resolution. We hypothesized that the phenotype of *Mtb*-specific T cells may be quantitatively impacted by the load of bacteria present in a patient. We recruited 105 Tanzanian adult tuberculosis (TB) patients and obtained blood before and after 5 months of antibiotic treatment. We studied relationships between patients’ clinical characteristics of disease severity and microbiological as well as molecular proxies of bacterial load in sputum at the time of diagnosis. Besides, we measured by flow cytometry the expression of CD38 or CD27 on CD4^+^ T cells producing interferon gamma (IFN-γ) and/or tumor necrosis factor-alpha (TNF-α) in response to a synthetic peptide pool covering the sequences of *Mtb* antigens ESAT-6, CFP-10 and TB10.4. Reflecting the difficulty to extrapolate bacterial burden from a single end-point read-out, we observed statistically significant, but weak, correlations between Xpert MTB/RIF, MBLA and time to culture positivity. Unlike CD27, the resolution of CD38 expression by antigen-specific T cells was observed readily following 5 months of antibiotic therapy. In addition, only the intensity of CD38-TAM signals measured at diagnosis significantly correlated with *Mtb* 16S RNA recovered from patients’ sputa. Altogether, our data support CD38-TAM as an accurate marker of infection resolution and a sputum-independent proxy of bacterial load.

## INTRODUCTION

Tuberculosis (TB) is an air-born infection caused by gram-positive bacilli called *Mycobacterium tuberculosis* (*Mtb*). In 2020, an estimated 9.9 million people fell ill of TB, of which 4.1 million were not diagnosed and 1.5 million died (1). As such, TB remains today the leading cause of death from a single infectious agent after the SARS-CoV-2 coronavirus (COVID-19). Yet, TB is mostly treatable with a combination of drugs for a minimum of 6 months. Unfortunately, side effects and hepatotoxicity in particular has been observed for up to 28% of patients (2). In addition, lung imaging by tomography coupled to the detection of short-lived mRNA species within sputa demonstrated that nearly a quarter of patients would still harbor active lesions and live mycobacteria in their lungs upon treatment completion (3). This may be linked to *Mtb*’s capacity to respond to immune-induced stresses by switching into a dormant state, metabolically associated with drug tolerance, as well as the limited penetration of antibiotics into caseous lung lesions and cavities (4, 5). Thus, despite being apparently cured, many patients may still harbor live bacilli in their lungs at the end of their treatment. TB control strategies should therefore not only rely on improving diagnosis and shortening treatment, but also monitoring bacterial clearance to ensure treatment efficacy and disease resolution (6, 7). One particular problem is that, without accurate monitoring tools, all TB patients have to complete 6 months of treatment despite evidence suggesting cure after 4 months in substantial proportion of patients (8), while others may develop drug-resistant (DR)-TB requiring adjusted treatment regimens. DR-TB treatment are longer, often poorly tolerated and have lower cure rates (9, 10). Shorter TB treatment regimens involving new formulations, new compounds or repurposed drugs are being tested in clinical trials to treat drug-sensitive as well as DR-TB (6). These trials are challenged by the difficulty to accurately quantify and compare treatment responses (11, 12). Sputum-based treatment monitoring tools such as smear microscopy, looking at acid-fast bacilli positivity conversion after two months of treatment initiation, have low sensitivity (13, 14). Moreover, this test cannot distinguish between dead and live bacilli and was shown to be poorly sensitive and specific of treatment failure or relapse (15). Molecular-based methods, such as Xpert MTB/RIF, cannot differentiate between live and dead bacteria either (16). *Mtb* sputum culture results require up to 8 weeks for solid culture and 42 days for liquid culture thus cannot be used to assess the patient’s response to treatment in a timely manner (17). Furthermore, collection of sputum samples beyond 2 months of treatment is challenging as patients’ coughs have often resolved by then (7). Early bacterial activity (EBA) assessing bacterial decline over 14 days following onset of treatment, has been implemented but this method cannot detect the eventual development of DR-TB (18, 19). Molecular methods such as quantitation of 16S rRNA using molecular bacterial load assay (MBLA) have been employed in early bacterial activity studies to reflect bacterial decline as early as 3 days following initiation of treatment, predict relapse, monitor treatment response and are closely correlated with culture-based read-outs (20, 21). However, extrapolating the bacterial burden of a patient from a single sputum sample is inherently inaccurate due to the stochastic release of bacteria across independent coughing episodes and the impossibility to sample bacilli outside the lung compartment in the case of extra-pulmonary TB. In that context, non-sputum-based monitoring tools may be more appropriate. Blood-based biomarker tests constitute promising tools for sputum-independent diagnosis of TB and, by extension, monitor treatment response and predict treatment outcome (7, 8, 22). IFN-γ and/or TNF-α-producing CD4^+^ T cells expressing activation, differentiation or proliferation markers have been repeatedly explored for TB diagnosis (7, 13, 17, 22-25). In addition to their diagnostic potential, these T cell activation markers (TAM) have been investigated for their ability to monitor bacterial clearance, disease extent and treatment outcome (22, 23, 26, 27). Compared to CD27 and Ki67, CD38 and HLA-DR expressed by *Mtb*-specific CD4^+^ T cells declined more rapidly following anti-TB treatment initiation (8, 17, 23). The frequencies of antigen-specific CD4^+^ T cells expressing HLA-DR or Ki67 have been associated with disease severity and time to culture conversion (23, 24, 28). In that context, we aimed to assess the correlation of CD38 and CD27 biomarkers with measures of disease severity including sputum bacterial load and in response to anti-TB treatment using a standardized whole-blood TAM assay.

## METHODS

### Study design and participants

We collected samples from 105 drug-sensitive TB patients prospectively recruited from the National Tuberculosis and Leprosy Program (NTLP) TB clinic, who were enrolled as a part of an ongoing TB-Dar cohort study (25). All consecutive adult TB patients (age ≥18 years) with a positive sputum smear microscopy (Ziehl-Neelsen staining) and/or *M*.*tuberculosis* detected by Xpert MTB/RIF were eligible for the study. Clinical assessment was done at the time of diagnosis as well as after 2-, 5- and 12-months to assess patient response to anti-TB treatment and relapses. Blood specimens were collected at the time of TB diagnosis and 5 months after anti-TB treatment initiation. In addition, patients provided sputum samples at the time of study enrolment and, when possible, at month 5 for assessment of treatment success. The study protocol was approved by the institutional review board of the Ifakara Health Institute (IHI; reference no. IHI/IRB/EXT/No: 16-2019) and the Medical Research Coordinating Committee of the National Institute for Medical Research (NIMR; reference no. NIMR/HQ/R.8a/Vol.IX/1641) in Tanzania. All participants provided a signed informed consent to collect clinical data, sputum and blood samples. In case of illiteracy, study information was given in the presence of an impartial, literate witness, who read the information sheet to the participant or witnessed the complete reading of the information sheet to the participant.

### TB score

Excepting tachycardia (data not collected), the TB score was adapted based on ten of the eleven signs and symptoms of TB disease that were previously reported as a potential measure of treatment outcome in resource-limited settings (29). The TB score used here therefore encompassed cough, hemoptysis, dyspnea, chest pain and night sweat self-reported symptoms as well as pallor anemia, positive finding at lung auscultation, fever (>37°C), body mass index (BMI) <18, BMI<16, middle upper arm circumference (MUAC) <220 and MUAC <200. Each parameter accounted for 1 point adding up to a maximum of 12 points.

### Microbiological procedures

The Xpert MTB/RIF test was performed at the NTLP laboratory and only positive patients were enrolled for this study. On the day of enrolment and prior initiation of treatment, an early morning sputum specimen was collected for culture and bacterial load assessment. The samples were first homogenized for 30 min then treated with cetylpyridinium chloride at 25°C for 4-7 days to increase culture recovery as reported previously (30). A final decontamination step with 1% NaOH (final concentration) was performed before culture on Lowenstein–Jensen media at 37°C. For molecular bacterial load assessment (MBLA), one ml of homogenized sputum was mixed with freshly thawed guanidine thiocyanate solution (4M GTC, Promega; V2791) in 1M Tris-HCl pH 7.5 complemented with 1% β-mercaptoethanol shortly before usage. GTC-treated sputa were left standing for 2 h at 25°C before storage at -80°C.

### RNA extraction and quantitation

Sputum samples in GTC were centrifuged at 3000 g for 30 minutes. Pellet was suspended in 500 μl of TRI Reagent® (Zymo; R2050-1-200) and stored at -80°C. TRI samples were spiked with 50 ng of internal control (IC) made of a 1957bp sequence of the potato gene phyB prepared as described elsewhere (31) and transferred to BeadBug tubes (SIGMA; Z763721). Subsequently, samples were bead-beaten using a FastPrep-24 (Life sciences) for 25 s at 6.5 m/s, cooled on ice for 5 min, bead-beaten for an additional 20 s cycle and placed back on ice. RNA extraction was performed using the Direct-zol™ RNA Microprep kit (Zymo; R2062) according to manufacturer’s instructions. DNA contamination was assessed following 16S quantitative PCR (qPCR) using HOT FirePol EvaGreen qPCR mix plus (Solis BioDyne: 08-24-0000S) on a StepOnePlus Real-Time PCR System (ThermoFisher). Samples with a 16S cycle threshold (Ct) value <35 were treated with RQ1 DNase (Promega; M6101) and purified using RNAeasy MinElute® cleanup kit (Qiagen; 74204) according to manufacturer’s protocol. Multiplex reverse transcription (RT)-qPCR was performed using SOLIScript 1-step Multiplex Probe Kit (Solis BioDyne; 08-59-0000S) and the primers and probes listed in supplementary table 1. Each sample was assessed in duplicate plus a minusRT control.

**Table 1:** Demographics and clinical characteristics before and after 5 months of treatment.

| Characteristics | Before treatment<br>(n=105) | 5 months after treatment<br>initiation | p value |
| --- | --- | --- | --- |
| Age in years, media (IQR) | 32 (25-40) | - |  |
| Age group in years, n (%) |  |  |  |
| 18-24 | 19 (18.1) | - |  |
| 25-34 | 44 (41.9) | - |  |
| 35-44 | 26 (24.8) | - |  |
| ≥45 | 16 (15.2) | - |  |
| Female, n (%) | 32 (30.5) |  |  |
| **BMI, kg/m<sup>2</sup>, median (IQR)** | **18.06 (16.87-19.71)** | **19.76 (17.86-21.13)** | **<0.001** |
| HIV<sup>+</sup>, n (%) | 12 (11.4) | 13 (12.64) |  |
| On ART, n (%) | 12 (100) | 13 (100) |  |
| Symptoms <sup>1</sup>, n (%) |  |  |  |
| Cough | 103 (98.1) | - |  |
| Positive finding at lung auscultation | 93 (88.6) | - |  |
| Fever | 74 (70.4) | 10 (9.5) |  |
| Chest pain | 72 (68.6) | - |  |
| Night sweat | 48 (45.7) | - |  |
| Hemoptysis | 11 (10.5) | - |  |
| Anemic conjunctivae | 6 (5.7) | - |  |
| Dyspnoea | - | - |  |
| BMI<18 | 51 (48.6) | 28 (26.7) |  |
| BMI <16 | 13 (12.4) | 2 (1.9) |  |
| MUAC <220 | 18 (9.5) | 12 (11.4) |  |
| MUAC <200 | 7 (6.7) | 1 (0.9) |  |
| TB patient category, n (%) |  |  |  |
| New | 101 (96.2) | - |  |
| Relapse | 4 (3.8) | - |  |
| Treatment after default | - | - |  |
| Treatment outcome cured | na | 105 (100) |  |
| Full blood counts (10<sup>9</sup> cells/L) |  |  |  |
| White blood cells, median (IQR) | 8.080 (6.310-9.750) | nd |  |
| Platelets, median (IQR) | 299 (242-400) | nd |  |
| Red blood cells, mean (±SD) | 4.841 (0.90) | nd |  |
| Xpert MTB/RIF Ct values results |  |  |  |
| Ct values median (IQR) | 19.40 (15.90-22.60) |  |  |
| HIV<sup>+</sup>, Ct values median (IQR) | 25.45 (18.85-28.18) |  |  |
| **HIV<sup>-</sup>, Ct values median (IQR)** | **19.10 (15.60-22.00)** |  | **0.04665<sup>§</sup>** |
| Smear results, n (%) |  |  |  |
| Scanty | - |  |  |
| 1<sup>+</sup> | 6 (5.7) | - |  |
| 2<sup>+</sup> | 5 (4.8) | - |  |
| 3<sup>+</sup> | 4 (3.8) | - |  |
| Negative | - | 75 (71.4) |  |
| Not done | 90 (85.7) | 30 (28.6) |  |

BMI, Body mass index; HIV, Human immunodeficiency virus; AFB, Acid-fast bacteria; MTB, *Mycobacterium tuberculosis*; ART, Antiretroviral therapy. na, not applicable; nd, not done; Ct, Cycle threshold; neg, negative. Smear results: Scanty, 1-9 AFB per 100 fields; 1+, 10-99 AFB per 100 fields; 2+, 1-10 AFB per field in at least 50 fields; 3+, more than 10 AFB per field in at least 20 fields; Negative, no AFB in at least 100 fields. § compared to HIV<sup>+</sup> arm.

### MBLA normalization

To account for potential RNA loss during the extraction and amplification inhibition, we assessed the correlation between RT-qPCR detection of 16S rRNA and the IC across 24 technical replicates of RNA extraction from an H37Rv culture (31). Linear regression was performed and the resulting slope was used for IC-based normalization of the 16S rRNA Ct value obtained for each sample (Supplementary material).

### MBLA standard curve

To establish the linearity and the limit of detection of the MBLA assay, triplicates of 10-fold serial dilutions were prepared from an exponentially growing H37Rv culture (OD 0.51). Bacterial concentration was quantified by CFU assessment on 7H11/OADC agar plates. 16S rRNA Ct values were normalized using the equation 16S rRNA Ct – [(IC Ct – 16.00) × slope] and plotted against the bacterial amount determined by CFU (ranging approximately from 5×10^5^ to 5 CFU/ml). Best-fit linear equation was subsequently used to extrapolate sputum bacterial load from the respective normalized 16S rRNA Ct value obtained from the sputum specimen (Supplementary material).

### TAM-TB assay

Freshly collected blood or cryopreserved PBMCs were processed for TAM-TB assessment as recently described (25). The following reagents were obtained through BEI Resources, NIAID, NIH: Peptide Array, *Mycobacterium tuberculosis* ESAT-6 Protein, NR-50711, *Mycobacterium tuberculosis* CFP-10 Protein, NR-50712 and *Mycobacterium tuberculosis* TB10.4, NR-34826. TAM-TB outputs in this study were made of ratios between the frequency of cytokine-producing CD4^+^ T cells expressing the investigated biomarker (CD27 or CD38) divided by the frequency of cytokine-producing CD4^+^ T cells negative for the respective biomarker of interest.

### Statistical analyses

Proportions and measures of central tendency (mean or median) were used to describe patient characteristics as detailed in Table 1. Mann-Whitney U test was performed using R version 4.0.3 to test differences in medians between groups. Analysis of variance (ANOVA), linear regression and Wilcoxon matched–paired rank tests were performed with GraphPad Prism 8.2.1.

## RESULTS

### Study cohort

Clinical data at the time of phlebotomy are summarized in Table 1. All study participants had drug-sensitive TB defined by Xpert MTB/RIF (Xpert) test results with a median age of 32 years [IQR: 25-40]. 12 patients (11.4%) were HIV co-infected, 11 (91.7%) of whom were already on antiretroviral treatment (ART) at the time of TB diagnosis and one initiated ART during the course of antibiotic therapy. Consistent with previous reports (32) and despite ART, HIV co-infected participants presented with a significantly lower bacterial load at the time of TB diagnosis with a reported median Ct value of 25.45 [IQR:18.85-28.18] compared to 19.40 [IQR: 15.60-22.00] in HIV-negative patients (p= 0.0467). 75 patients (71.4%) were still able to produce sputum specimen after 5-months of anti-TB treatment (ATT) of which all were found smear negative by microscopy. TB clinical severity reflected by TB score showed a significant resolution of disease (p<0.001) after 5-months of ATT (Figure 1E). Excepting mild fever episodes (<37.8°C) in 9.5% of patients, major TB symptoms had resolved in all participants after 5 months of ATT yet, anti-TB treatment was pursued for another month as per NTLP guidelines (33). Consistent with the complex reverse causality between weight loss and undernutrition in TB pathogenesis and susceptibility (34), we observed a significant increase in median BMI (18.06 kg/m^2^ [IQR: 16.87-19.71] to 19.76 kg/m^2^ [IQR: 17.86-21.13], p=0.001) after 5 months of TB treatment.

**Figure 1.**
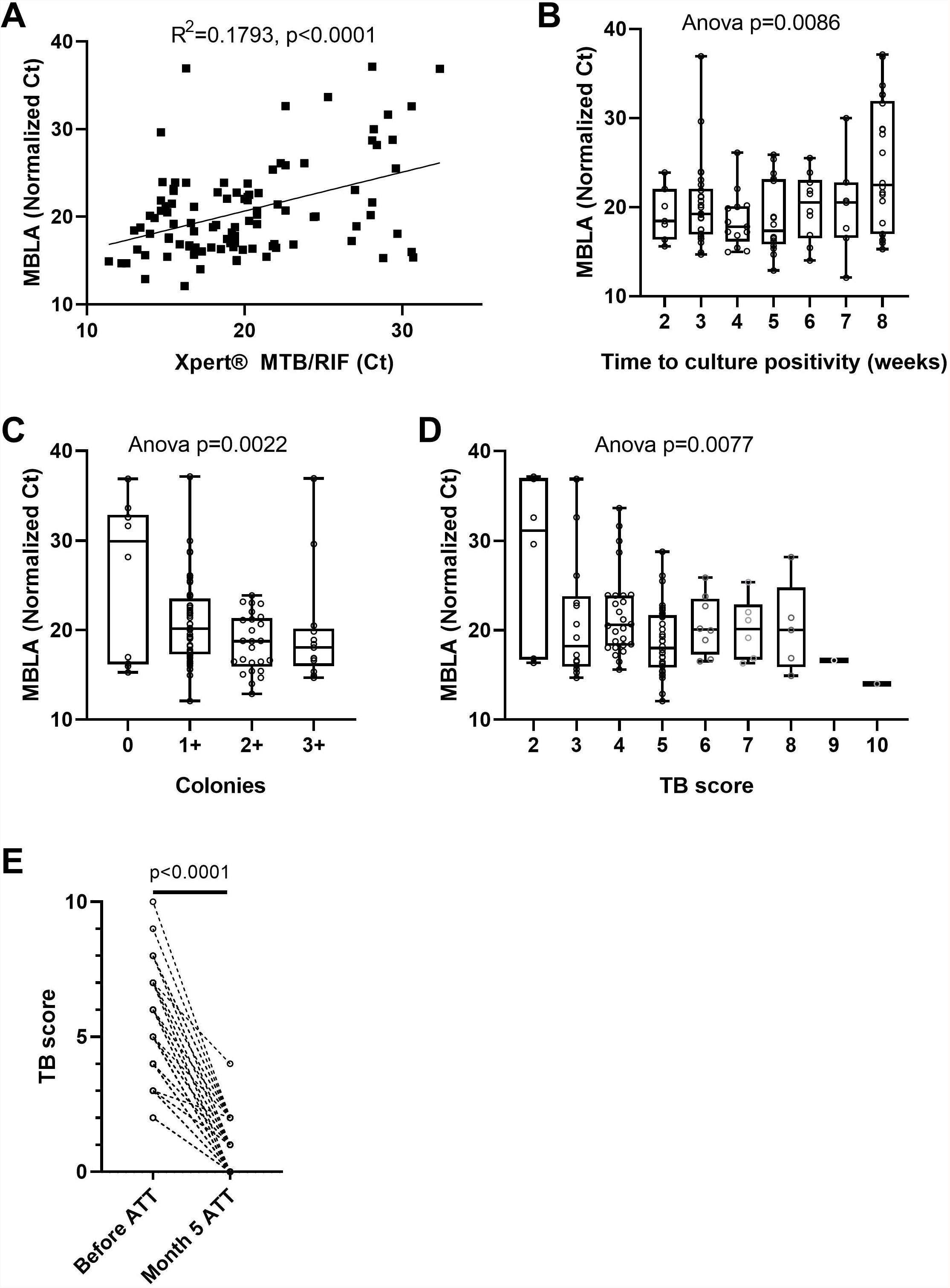
Concordance between various estimates of sputum bacterial load and clinical assessment of disease severity across TB patients at time of diagnosis and after 5 months of antibiotic treatment. Sputum molecular bacterial load assay (MBLA) results against: A) Xpert MTB/RIF Ct values results from an independent sputum specimen or B) time to culture positivity in weeks, or C) solid culture intensity grades from the same sputum specimen and D) patient’s clinical TB score at time of diagnosis. E) Evolution of clinical TB score after 5 months of anti-tuberculous treatment (Month 5 ATT). A) Pearson correlation p-value (two-tailed) and R squared. B), C) and D) One-way ANOVA p-values. E) Wilcoxon matched-pairs signed rank test p-value.

### Sputum bacterial load and TB disease severity

To counteract the poor discriminative value of Xpert likely originating from its capacity to detect cell-free DNA and DNA from dead bacteria (35), we used the 16S rRNA-based molecular bacterial load assay (MBLA) (31) to estimate the number of viable *Mtb* bacilli present in sputum at baseline. MBLA Ct values plotted against Xpert Ct values revealed a statistically significant, yet moderate, correlation between the two measurements (Figure 1A). We then investigated the relationship between the MBLA results and the readouts from solid culture-based encompassing time to culture positivity (TTP) (Figure 1B) and colony counts (Figure 1C). Lower MBLA Ct values were associated with shorter TTP and higher bacterial counts, yet substantial discrepancies remained. For instance, sputum samples producing similar MBLA Ct values also displayed different TTP or colony counts. Finally, we sought to explore the relationship between sputum bacterial load and clinical disease severity reflected by the TB score, which accounts for the presence of self-reported symptoms as well as other relevant clinical parameters (29). While we could observe a trend for extremely low or high clinical scores associating with rather high and low MBLA Ct respectively, patients with a TB score ranging between 3 and 8 displayed highly overlapping bacterial load (Figure 1D). Together, our data suggest that bacterial load estimates relying on a single end-point sputum specimen are unlikely to accurately reflect the absolute amount of bacteria present in a patient. Hence, we next determined the accuracy of the non-sputum-based TAM-TB assay to discriminate between active and cured TB patients and whether the intensity of the TAM-TB signals correlate with MBLA Ct values.

### CD38-based TAM-TB accurately reflects *Mtb* clearance following antibiotic treatment

We recently reported the accuracy of CD27-and CD38-based TAM-TB assay to diagnose TB in a cohort of adults with presumptive TB (25). In the current study, we aimed to compare the evolution of CD27 and CD38 upon bacterial clearance following antibiotic treatment by performing the assay on blood collected at the time of TB diagnosis prior treatment initiation and after 5 months of anti-tuberculous treatment and clinical confirmation of disease resolution. We ran the CD27-and CD38-based TAM assays side-by-side. The frequency of cytokine-producing T cells across the two independent stimulations for all collected specimens is plotted in Figure 2A to demonstrate the robustness of the assay to recall *Mtb*-specific T cell responses in a reproducible manner (slope = 1.043, R^2^= 0.9299). Overall, we observed a significant drop in the frequency of antigen-specific T cells recalled by the synthetic peptide pool after 5 months of treatment (Figure 2B; median response before and 5 months post-ATT: 0.131% and 0.0773% respectively, p<0.001). Nonetheless and consistent with the concurrent resolution of TB with the effector phenotype of specific T cells, the expression of CD27 and CD38 was significantly increased or decreased respectively during ATT (Figure 2C-D). Moreover, a receiver operating characteristic (ROC) curve analysis revealed a substantially superior resolution of CD38 compared to CD27 with an area under the ROC (AUROC) of 0.953 versus 0.758 (Figure 2E).

**Figure 2.**
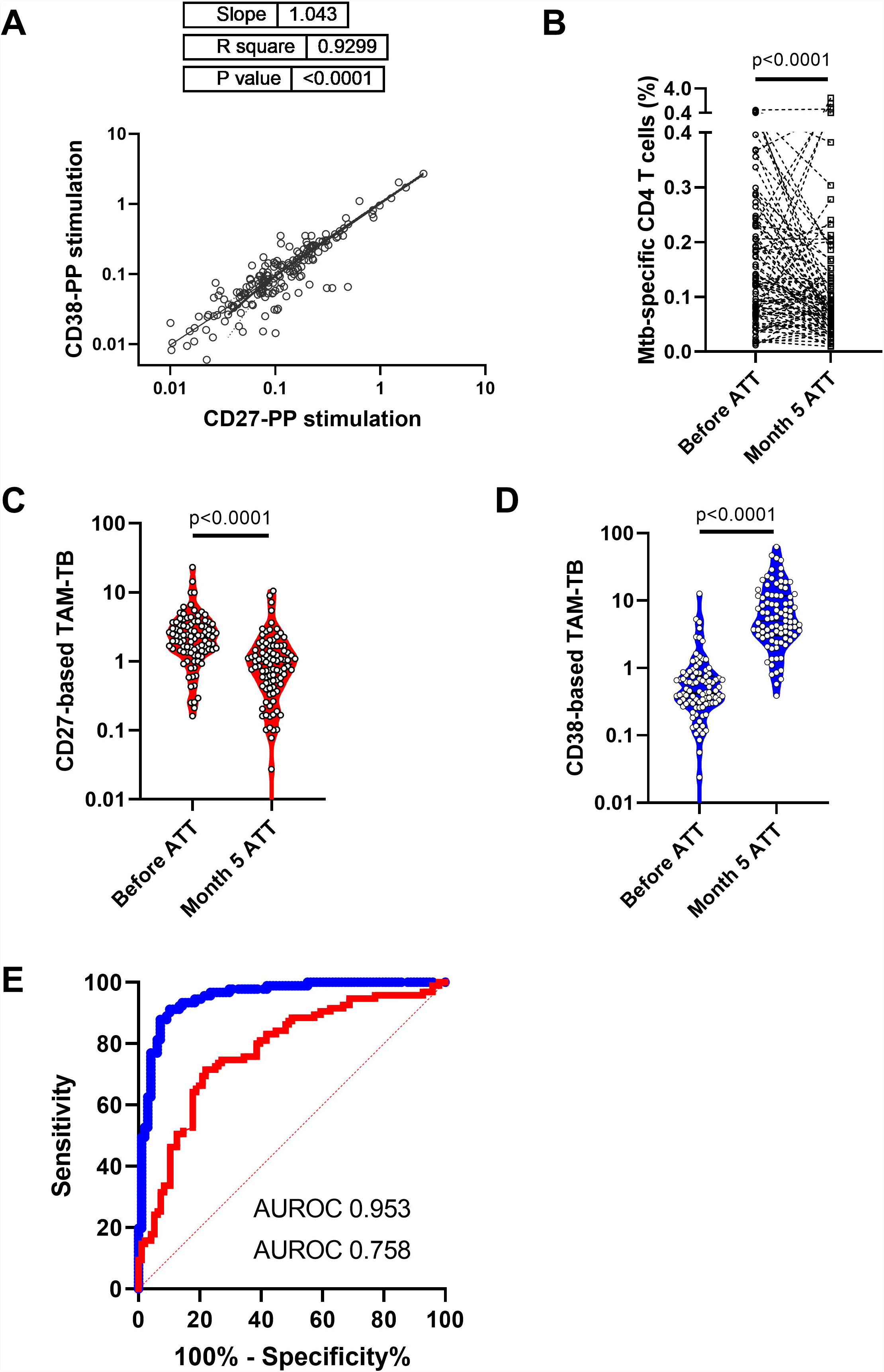
Superiority of CD38-over CD27-based T cell activation marker to attest TB infection resolution following 5 months of anti-tuberculous treatment. A) Absolute frequencies of *Mtb*-specific CD4 T cells producing cytokines across independent stimulation of peripheral blood mononuclear cells specimen from the same patient and study visit. B) Median response of the frequencies of antigen-specific T cells recalled by the synthetic peptide pool before or after 5 months of anti-tuberculous treatment (Month 5 ATT). C) Evolution of T cell activation markers expressed by *Mtb*-specific CD4 T cell before and after 5 months of antibiotic treatment. D) Receiver operating characteristic (ROC) curve and area under the ROC curve showing the superior discriminatory power of CD38-over CD27 biomarkers to discriminate *Mtb*-specific CD4 T cell responses of patients before and after 5 months of chemotherapy.

### CD38-based TAM-TB constitutes a sputum-independent proxy of bacterial load

Since TAM-TB signals were found to be actively promoted by ongoing infection, we next studied whether their intensity may be modulated in a quantitative manner by the amount of bacteria detected via MBLA within the same patient. We found that the CD27-based TAM-TB intensities did not decrease with the amount of mycobacterial RNA recovered in the sputum of the respective patient (Figure 3A). In contrast, we observed a rather poor goodness of fit, yet, a highly significant deviation from a non-zero slope between MBLA Ct values and the decline of CD38 positive antigen-specific CD4^+^ T cells that translate into increasing CD38-based TAM-TB results when expressed as a ratio with CD38 negative antigen-specific CD4 T cells (Figure 3B).

**Figure 3.**
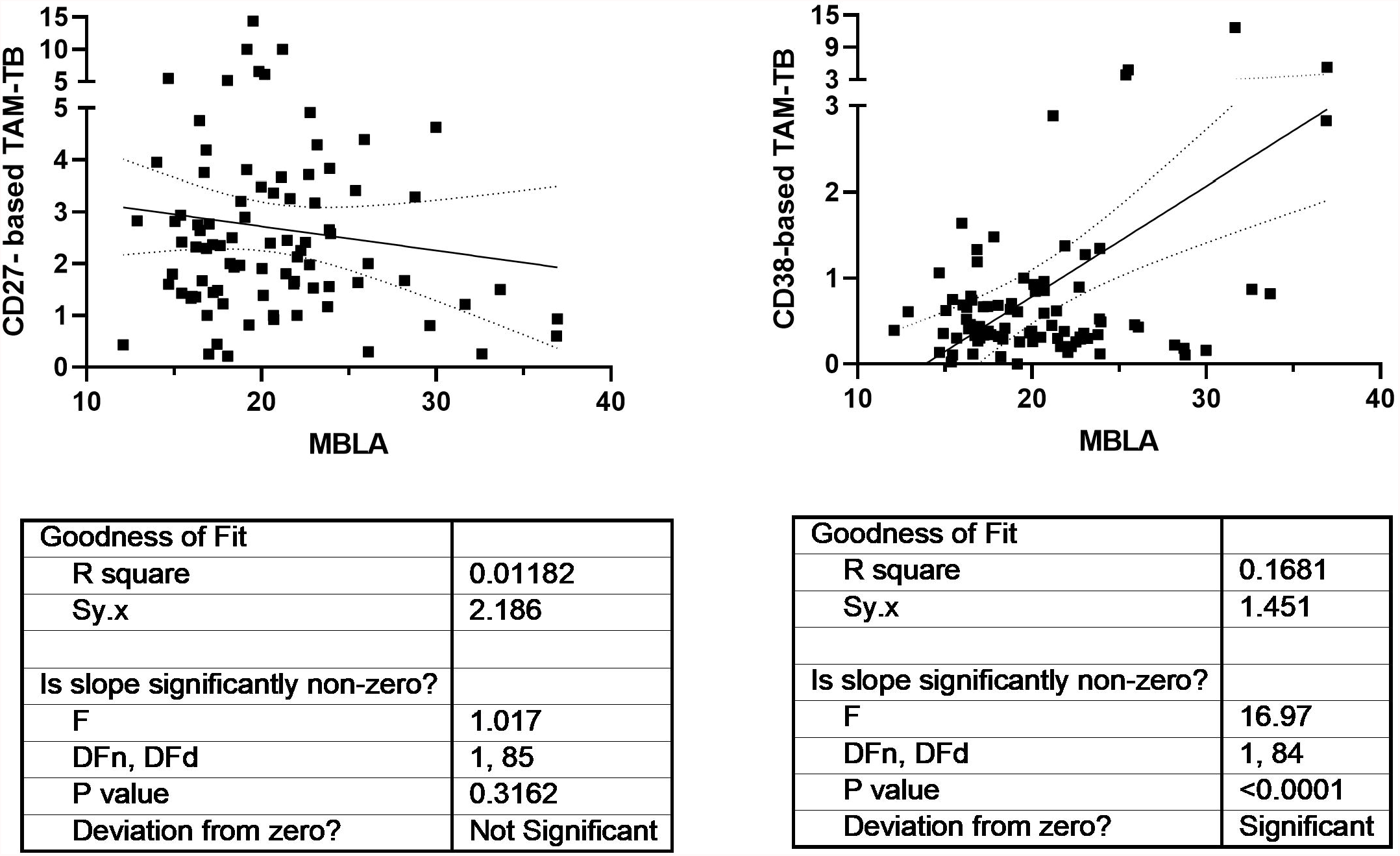
Superiority of CD38 -over CD27-based T cell activation marker (TAM) to correlate with sputum bacterial load before anti-tuberculous treatment initiation. Linear regression analysis comparing the strength of the association between CD27-or CD38-biomarker expression by *Mtb*-specific CD4 T cells (CD27-or CD38-based TAM-TB) and sputum molecular bacterial load assay (MBLA) results from respectively, blood and sputum specimen of TB patients at time of diagnosis.

## DISCUSSION

We hypothesized that during an infection, the bacterial antigens drained into secondary lymphoid organs trigger or recall T cell activation in an antigen-specific manner, and that the effector phenotype intensity of the *Mtb*-specific T cell population that recirculate in the periphery may therefore reflect the bacterial load independently of the infection site. This assumption implies that TAM signals should wane upon bacterial clearance by ATT and that they should also display signs of correlation with alternative read-outs of bacterial load. Side-by-side evaluation of CD27-versus CD38-based assays to distinguish active from cured TB were previously reported in a pilot study and as a case report in the context of extra-pulmonary TB (8, 17). Our study benefited from a highly systematic comparison and an unprecedented sample size. In that context, we could demonstrate the superiority of CD38 to resolve swiftly after 5 months of ATT and concomitant disease resolution (Figure 2E). This observation is consistent with the studies from Halliday *et al*. and Streitz *et al*., that reported the incapacity of CD27 to distinguish between recently and remotely acquired latent TB infection (LTBI) and the very slow reversion of CD27 downregulation in patients up to one year after therapy (36, 37). Our results also concur with the excellent performance of CD38 to diagnose TB with no interferences arising from LTBI (13, 25). The superior capacity of CD38 to monitor treatment responses is also consistent with the data presented here demonstrating the incapacity of CD27-compared to the unique propensity of CD38-based TAM-TB assay to be influenced by the amount of live bacteria recovered from patient sputum. This difference suggests a differential expression kinetics for each marker upon T cell priming or recall and/or that a differential threshold of antigen concentration may be required to stimulate or maintain the expression of the two markers. Another T cell marker, CD153, has been recently reported to correlate with bacterial load in non-human primate models as well as in TB patients (22). Conversely, ATT only partially restored CD153 expression in the *Mtb*-specific CD4 response. In the absence of a gold standard, we cannot attest the superiority of our immunodiagnostic tool over microbiological and molecular approaches to assess *Mtb* bacterial burden most accurately. Previously, sputum bacterial load assessment determined by colony forming units or culture time to positivity correlated only moderately with molecular detection methods based on DNA such as Xpert MTB/RIF assay (35). Consequently, we implemented the recently developed 16S rRNA-based molecular bacterial load assay (MBLA) (31) to estimate viable *Mtb* bacilli within the collected sputum specimens. As expected, we observed a moderate correlation between DNA-and RNA-based read-outs across the cohort of TB patients. Specimens displaying high MBLA Ct values despite low Xpert Ct values could be readily explained by the detection of cell-free DNA or DNA from dead bacteria. However, the reciprocal scenarios are more difficult to reconcile. Such discrepancies between the two molecular methods could first have arisen from sampling variability. Indeed, Xpert results were provided by the NTLP laboratory while MBLA tests were performed on independent different sputum sample collected on the day of study enrolment. Alternatively and non-exclusively, a high prevalence of dormant bacteria (38) harboring limited transcriptional activity could also fuel such differences. We also found discrepancies between MBLA Ct values and culture-based read-outs of sputum bacterial load. In that regard, differentially cultivable mycobacteria across liquid and solid media, as well as bacterial subpopulations requiring resuscitation factors to grow, may affect bacterial recovery (39, 40). We observed a rather poor relationship between the amount of bacteria recovered from sputum specimens collected at the time of TB diagnosis and clinical disease severity scores (Figure 1D). Since TB score has been found to significantly and inversely correlate with survival estimates, our data suggest that TB disease severity does not always corresponds with greater bacillary burden. Actually, TB in HIV-infected individuals is generally associated with a lower bacterial load in the sputum but also poor prognosis (41). In addition, a single end-point measurement from a unique sputum specimen is unlikely to reflect accurately the absolute amount of bacteria present in the entire lung compartment. Furthermore, sputum-based read-outs may underestimate the bacterial burden of a particular host/strain combination that would favor the dissemination of the bacteria outside the lungs (42).

In conclusion, we observed a concomitant superiority of CD38 to discriminate active TB patients from cured individuals, as well as a significant capacity to be influenced by the amount of live mycobacteria that could be retrieved from the patient’s sputa. Taken together, our results suggest that the expression of the T cell activation marker CD38 is induced quantitatively by the presence of live mycobacteria during TB. As such, the CD38-based TAM-TB assay does not only constitute an excellent assay to monitor treatment response but also a good candidate for the assessment of the bacterial burden within, but also beyond, the lung compartment. In that context, we believe that the CD38-based TAM-TB assay is a promising avenue to diagnose asymptomatic subclinical forms of TB that are urgently required in TB control strategies to end TB (43).

## Supporting information

Supplementary material_Hiza H et al

## Data Availability

All data produced in the present study are available upon reasonable request to the authors

## Acknowledgements

We would like to express our sincere gratitude to Prof. Timothy D McHugh and his team at UCL Centre for Clinical Microbiology for providing initial training and the IC plasmid for the implementation of the MBLA in our institutes.

