## Supplementary material_Hiza H et al for "CD38 expression by antigen-specific CD4 T cells correlates with sputum bacterial load at time of tuberculosis diagnosis and is significantly restored 5-months after treatment initiation"

**Supplementary table**

| **Primer** | **Covering sequence** |
| --- | --- |
| 16S-Fw | 5'-GTGATCTGCCCTGCACTTC-3' |
| 16S-Rv | 5'-ATCCCACACCGCTAAAGCG-3' |
| IC-Fw | 5'-GCACAGGGTTGATGTTGGTATTGTC-3' |
| IC-Rv | 5'-CAAATGAGAAATAGCCCTCACTGCAAG-3' |
| **Probes** | **Reporters, covering sequences and quencher** |
| 16S-FAM | 5'6FAM-AGGACCACGGGATGCATGTCTTGT-3'BHQ1 |
| IC-JOE | 5'-JOE-GCAGGGTCCTCAGTTCTAGCAGGCTCCA-3'BHQ1 |

**Supplementary figure**

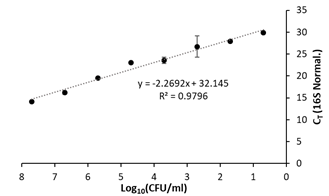
**A	B**

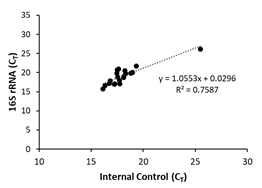

A) Linear regression analysis between cycle threshold (Ct) values of 16S rRNA of an H37Rv culture and the spiked internal control across 24 extraction/amplification replicates resulted in a slope of 1.0553. B) Linear regression analysis between IC-normalized 16S rRNA Ct values and bacterial concentration for an H37Rv dilution series (ranging from 5×10^7^ to 5 CFU/ml) processed in triplicate. The computed equation displayed on the plot was used to interpolate the bacterial load within the investigated sputum specimens from the normalized MBLA Ct values. CFU, colony-forming units.
